# Identifying Distinct Tourette Disorder Subtypes using Clinical Data

**DOI:** 10.1101/2025.11.09.25339700

**Authors:** Subramanian Krishnamurthy, Tourette International Collaborative Genetics (TIC Genetics), Robert A. King, Jay A. Tischfield, Gary A. Heiman, Jinchuan Xing

**Author notes:** Correspondence: Gary A. Heiman, Ph.D., Department of Genetics, Rutgers, The State University of New Jersey Nelson Biology Laboratories B412, 604 Allison Rd, Piscataway, NJ 08854, Jinchuan Xing, Ph.D., Department of Genetics, Rutgers, The State University of New Jersey Life Science Building 225, 145 Bevier Road, Piscataway, NJ 08854.

## Abstract

Tourette Disorder (TD), or Tourette Syndrome, is a highly heterogeneous childhood-onset neurodevelopmental disorder with a global prevalence of 0.5%. TD’s large phenotypic heterogeneity, variable heritability patterns, and frequent but varied comorbidity with other neurodevelopmental disorders indicate that different TD patients might have different etiologies. In this study, we performed unsupervised clustering of clinical data, such as categorical diagnoses and comorbidities, from 865 subjects with TD from the Tourette International Collaborative Genetics (TIC Genetics) study to detect TD phenotypic clusters. Using two different clustering methods, K-Means and Bayesian Hierarchical Clustering, we identified five distinct, clinically relevant phenotypic clusters. These phenotypic clusters are characterized by both previously described TD comorbidities, such as Obsessive-Compulsive Disorder (OCD) and Attention Deficit/Hyperactivity Disorder (ADHD), as well as other characteristics, such as sex and region. Defining clinically relevant TD phenotypic clusters could enable better diagnostics, treatment, and the understanding of TD etiology. In addition, stratified analysis of genetic data based on these phenotypic subtypes may help identify genes contributing to each TD subtype and provide insights into the disease variability.

## Introduction

Tourette Disorder (TD), also called Tourette Syndrome, is characterized by multiple motor tics and at least one vocal tic, occurring many times a day (usually in bouts) nearly daily or intermittently for at least a year, with an onset before age 18 [1]. Most subjects with TD over the age of 10 report premonitory urges that are relieved by tics [2]. Coprophenomena (such as copropraxia or coprolalia) may occur when tics are most severe [2]. In addition, there are also sex-dependent differences in the phenotypic expression of TD, with males more often showing tic-related behaviors and females more often exhibiting Obsessive-Compulsive Symptoms/Behaviors (OCS/OCB) [3, 4]. Twin and family studies have established that TD bears a strong genetic component with an unclear inheritance pattern, probably due to polygenic cause [5, 6]. Large-scale genetic studies of TD have identified specific classes of genetic variation that contribute to the genetic risk of TD, including common variants [7], segregating variants, rare copy number variants, and *de novo* variants that impact protein-coding sequences [8–11].

Approximately 88% of TD subjects have one or more other Neurodevelopmental Disorders (NDDs) as comorbidities [12], such as Obsessive-Compulsive Disorder (OCD, ∼30-50%), Attention Deficit/Hyperactivity Disorder (ADHD, ∼50-60%) and to a lesser extent, Autism Spectrum Disorder (ASD, ∼10%), as well as anxiety, depression, and other conditions [13]. Given the substantial genetic and phenotypic heterogeneity of TD, identifying distinct subtypes could assist in better diagnosis and treatment, as well as enable a better understanding of genetic architecture through stratified analysis of genetic data.

Several prior studies, reviewed in [14, 15], have attempted to identify common factors and subtypes of TD. Cavanna et al. [14] described eight studies of TD symptom clusters. Seven of these studies used symptoms (tics only, tics + complex tics, or tics + behavioral symptoms) to identify subtypes [16–22]. One study was based on the ADHD and OCD comorbidity of 952 subjects from 222 families [23]. A more recent review [15] identified 16 publications that published data-driven analysis of tic symptoms, behavioral studies and gene association studies to identify TD subtypes. In addition to studies described in [24], three new studies [9, 25, 26] performed psychometric subtyping based on tic symptoms and behavioral characteristics, and five recent studies [27–31] performed psychometric subtyping based on comorbidity features.

Although these studies provided useful information, they had limited ability to comprehensively identify TD subtypes, because many of them were based on cohorts with either limited subsets of available phenotypic information (i.e., comorbidities or symptoms), small sample size, and/or subjects that were demographically homogeneous. In addition, previous studies primarily used factor analysis or latent-class analysis methods such as Principal Component Factor Analysis (PCFA), Hierarchical Cluster Analysis (HCA), Latent Class Analysis (LCA), and Exploratory Factor Analysis (EFA). To our knowledge no study to date applies hypothesis-free unsupervised clustering methods to identify TD subtypes. In this study, we identified subsets with TD using unsupervised clustering of clinical and demographic data from the Tourette International Collaborative Genetics (TIC Genetics) study [32], a large international study with over 20 sites across the United States, Europe, Israel, and South Korea using two independent methods. This large, uniformly assessed clinical dataset enabled us to identify five phenotypic clusters that will enhance the understanding of the heterogeneity of TD.

## Materials and Methods

### Clinical Data

The clinical data were collected between 2011 and 2023 as a part of the TIC Genetics Study [32]. The Institutional Review Board approved the study protocol at each local site. Informed consent was obtained from all participants (or in the case of minors, from their parents). The clinical assessment methods and definitions of TD, OCD, ADHD, and Trichotillomania have been previously described in detail [32], and are based on the Diagnostic and Statistical Manual of Mental Disorders—Fourth edition, Text Revision (DSM-IV-TR) or Fifth edition (DSM-5) [1]. Clinicians were trained to record all symptoms and perform diagnoses in a consistent manner to ensure quality and reliability across all sites.

Detailed clinical data were acquired from an extensive written questionnaire which was completed by the participant or by the parent in the case of young probands. This information was validated in a semi-structured interview by an experienced clinician and recorded in a standardized fashion. Items included demographic information, details of tic, OCS/OCB/OCD, ADHD, and trichotillomania (including ages of onset), and other potentially relevant conditions. Based on the questionnaire and interview information, the clinician made lifetime diagnoses for 4,435 subjects, including probands with TD, both affected and unaffected parents, and some affected and unaffected siblings [32].

### Data Preprocessing

To improve the precision of subtype identification, we focused strictly on subjects with TD diagnosis and excluded other tic disorders, such as other chronic, provisional, and transient forms. Among subjects that had TD, we excluded those that have any missing or “unable to rate” value for any of the Tic, Obsessive Compulsive, or Attention-disorder-Hyperactivity diagnoses. We also excluded subjects with potentially confounding factors flagged by clinicians, including “atypical presentation”, “psychosis”, “other severe neurological condition”, “congenital anomalies”, “genetic syndrome/chromosomal abnormality”, “other significant psychiatric history”, and “other significant medical history”. From each family, we selected only one member diagnosed with TD to minimize confounding effects of shared genetics. Twenty-two centers around the world were grouped into 3 regions: USA, Europe, and Asia. A description of the detailed clinical data variables for each subject included in this study is available in Supplementary Table S1.

Some variables in our clinical dataset are mutually exclusive. For example, for OCD diagnosis, each subject belongs to one of the following categories: No OC disorder/symptoms, OC Subclinical, OC Symptoms, or OCD. In these cases, these mutually exclusive variables were combined into a single categorical variable (Supplementary Table S2). Because the age of onset for a given disorder was usually from parents’ recollection and the exact age has a high uncertainty, it was converted into two categories: “Early” (<= 10y) and “Late” (> 10y) based on the guidance from clinicians. In a few cases (<10%) where the age of onset was not available, age at diagnosis was used as a proxy. The age of onset of subjects with no diagnosis of the disorder was coded as “Never”. We used two categories for birth: “single” or “multiple” (>=2). After preprocessing, 14 categorical variables were selected for clustering (Supplementary Table S2).

### Exploratory data analysis and clustering method testing

Characteristics and distribution of individual variables were analyzed to examine the dataset. Pairwise associations between variables were calculated as Cramer’s V statistics using the “CramersV” function in the R package “rcompanion” (version 2.5) [33]. Several clustering methods were tested (Supplementary Table S3), and two unsupervised clustering methods, K-Means and Bayesian Hierarchical Clustering (BHC) [34], were selected to identify TD subtypes.

### K-Means clustering

K-Means clustering is an unsupervised clustering method that partitions n observations into k clusters in which each observation belongs to the cluster with the nearest cluster mean. Multiple Correspondence Analysis (MCA) [35] was used to convert categorical variables into principal components (PCs). This enabled us to: 1. reduce noise by removing less significant PCs with small contribution to the overall variance, and 2. use clustering methods that require continuous variables. K-Means clustering was performed using the K-Means function in R using the “stats” package (version 4.4.3). The number of clusters (centers = k) from 2 to 20 were evaluated. To ensure robust convergence and reproducibility, the algorithm was run with 5,000 iterations and 1,000 random starts.

Euclidean distance was used as the distance metric and the clustering method described by Hartigan and Wong [36]. The optimal number of clusters was identified based on the silhouette statistics, which combines cluster cohesion (how well each data point fits within its assigned cluster) and cluster separation (compared to other clusters) into a single, interpretable value.

### Bayesian Hierarchical Clustering (BHC)

The BHC method performs bottom-up hierarchical clustering, using a Dirichlet Process (infinite mixture) to model uncertainty in the data and Bayesian model selection, to identify clusters that can be merged at each step [34]. This method works with categorical data and automatically optimizes Dirichlet process concentration variable to infer the optimal number of clusters. BHC clustering was performed using the BHC BioConductor package (version 1.56.0) in R with default variables.

### Cluster characteristics

Variable importance was inferred using V-measure (or Normalized Mutual Information, calculated by “V-test” using “FactoMineR”, version 2.11, in R). V-measure is a single metric, computed based on percentage of the cluster that has a given characteristic and percentage of the population with a given characteristic that is in the cluster, which is an average of homogeneity and completeness. Key variables that define the cluster characteristics have high V-measure. The relative importance of each variable on each cluster was determined by Fisher’s exact test or V-Test.

### Tools

R was used for all statistical analyses, and a list of all R packages used in the analysis is in Supplementary Table S4.

### Data availability

All data used in this study are provided in Supplementary Table S1.

### Code availability

All codes used in this study are available on Zenodo at 10.5281/zenodo.21812294.

## Results

### Data pre-processing and exploratory analyses

An overview of the data processing and analysis pipeline is provided in Figure 1. For clustering, we selected subjects diagnosed with TD (1,548) and excluded subjects with genetic abnormalities or severe psychiatric conditions (421) (see Methods for details). In addition, we excluded subjects with who were rated “Unable to rate” for Tic, Obsessive-compulsive, or Attention-deficit-Hyperactivity disorder diagnoses. To limit the effects of genetic relatedness, we selected one offspring from each family. Subjects with missing data were filtered out. The final data set used for identifying subtypes includes 865 offspring diagnosed with TD. Counts by category for each variable are included in Supplementary Table S2.

**Figure 1.**
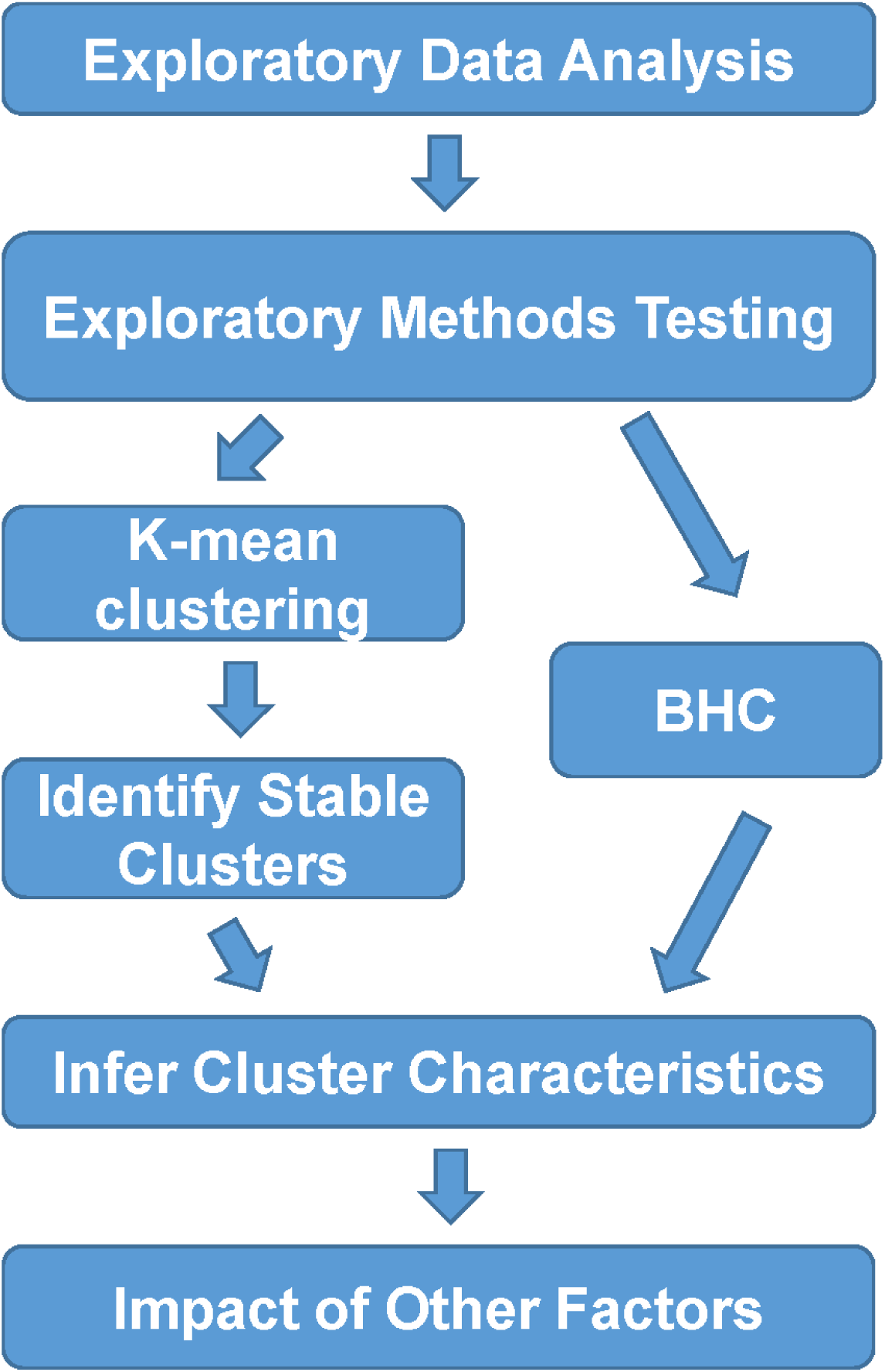
Analysis Pipeline overview. We first performed exploratory data analysis to understand the characteristics of data, such as variable distribution, completeness, and correlation. We then evaluated several unsupervised clustering methods (Supplementary Table S3) to identify methods that produce stable clusters. After testing, we selected two methods, K-Means and BHC for the final clustering and analysis of the data. After the cluster identification, we used V-Test to identify key factors that define the characteristics of each cluster and Fisher’s exact test to assess the impact of factors on each cluster.

To understand the relationship between selected variables, we computed pairwise association among the variables using Cramer’s V statistic (Supplementary Figure S1). The observed high association between OCD-related variables, ADHD-related variables, and Trichotillomania-related variables are expected. There were no other significant associations among the variables.

After evaluating eight unsupervised clustering methods (Supplementary Table S3), and many different subsets of data (Supplementary Table S5), we selected two methods that produced stable clusters with our dataset: K-Means (after transforming the data using MCA) and Bayesian Hierarchical Clustering. Both K-Means clustering after MCA [37] and BHC [38] mitigate the effects of correlation between variables. Therefore, we retained the correlated variables to provide additional insight into specific cluster characteristics.

### K-Means Clustering

To perform K-Means clustering, we used MCA to transform the categorical data from the dataset into PCs (Supplementary Table S6). Scree plot shows the contribution of each PC to the overall variance (Supplementary Figure S2). When we provided all PCs to K-Means clustering in our initial testing, it resulted in unstable clusters. These results are probably due to inherent variability in the clinical data. The later PCs capture mostly noise rather than meaningful structure in the clinical data. Therefore, we evaluated systematically excluding the least important PCs, one at a time, to reduce the noise in the dataset. For each set of PCs, we assessed the cluster number (K) from 2 to 20 and repeated the analysis 100 times to assess the stability (Supplementary Figure S3). This resulted in the selection of the top three PCs (account for >35% of the variance), that produced seven stable clusters (Supplementary Figure 3). Although K=7 has the highest Silhouette width, K=5 also shows similar Silhouette width with fewer clusters (Silhouette Plot, Supplementary Figure 4). Therefore, we examined the characteristics of both 5 and 7 clusters.

First, we identified the variables that most significantly correlated (positively or negatively) with cluster identity at K=5 (Figure 2A). For example, trichotillomania diagnosis is positively correlated only with cluster 2 (TrichDiag=TRUE), indicating grouping of subjects with trichotillomania in this cluster. ADHD is negatively correlated with cluster 3 (ADHDDiag=No ADHD). Plotting cluster memberships along the top three PCs suggests robust clustering (Figures 2B–D).

**Figure 2.**
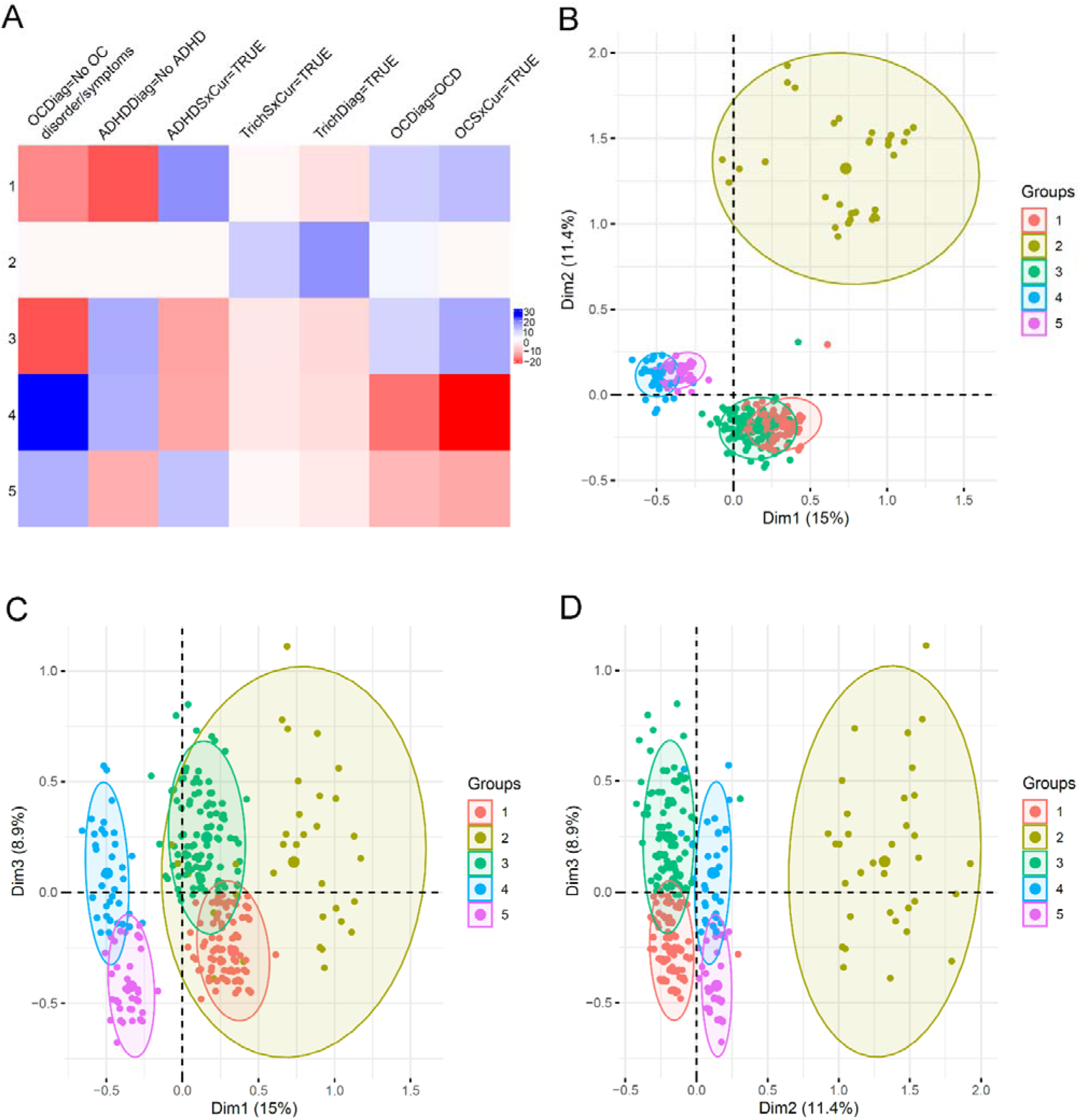
K-Means Clusters with K=5. **A.** Major factors determining cluster characteristics based on V-test scores. Positive values are positive correlations, while negative values are negative correlations. The larger the value, the stronger the correlation. Only statistically significant variables are plotted. See Supplementary Table S2 for detailed variable descriptions. **B-D.** Cluster plots on first three PCs. Each subject is shown as a dot, while the color of the dot corresponding to the cluster assignment. The centroid of each cluster is shown as a big dot. Ovals indicate 95% normal confidence regions around clusters’ 2D point cloud.

The detailed contribution and significance of each variable to each cluster are listed in Supplementary Data: KMeans_3Dim_5Clusters.xlsx, including percentage of subjects in a category in each cluster, percentage of subjects in each cluster in a category, percentage of subjects in a category in the whole data set, and v-test for the significance of each variable. These results provide details for each statistically significant variable for each cluster, allowing us to fully understand the cluster characteristics. Overall, 12 out of 14 variables show statistically significant contribution to the clusters, including all variables associated with OCD, ADHD, trichotillomania, region, and sex (Supplementary Data: KMeans_3Dim_5Clusters.xlsx– test.ch2 tab).

Based on the variable contribution to the clusters and the individual membership of each cluster, we determined the defining characteristics of each cluster (Supplementary Table S7). To make the results more comprehensible, we describe all diagnosis variables for a given disorder as one category in the sections below (e.g., “ADHD” for “ADHD diagnosis” and “ADHD current symptom”) and included the p-value for the most statistically significant variable.

Cluster 1 subjects (n=210) have ADHD (p=3.6E^-71^), early OCD age of onset (p=1.7E^-46^), no trichotillomania (p=1.1E^-5^), higher prevalence of males (p=2.2E^-8^) and ASD (p=7.9E^-4^). Cluster 2 subjects (n=40) have trichotillomania (p=6.7E^-70^), with higher prevalence of subjects from the USA sites (p=6.9E^-4^) and OCD (p=8.0E^-4^). Cluster 3 subjects (n=289) have high prevalence of OCD (p=4.0E^-52^), low levels of ADHD (p=4.0E^-49^), no trichotillomania (p=5.4E^-8^), higher prevalence of females (p=1.1E^-6^) and European subjects (p=1.8E^-3^). Cluster 4 subjects (n=229) have low NDD comorbidities, with no OCD (p=7.4E^-127^), very low ADHD (p=7.4E^-45^), no trichotillomania (p=3.3E^-6^), low prevalence of ASD (p=5.1E^-6^), and a high proportion of South Korean subjects (p=2.3E^-6^). Cluster 5 (n=97) comprises subjects with TD that have no OCD (p=5.6E^-45^), high prevalence of ADHD (p=2.6E^-32^), ASD (p=1.0E^-3^), and males (p=1.1E^-4^).

Next, we examined the contribution and significance of each variable to each cluster for K=7 (Supplementary Table S8, Supplementary Data: KMeans_3Dim_7Clusters.xlsx). Overall, 13 out of 14 variables show statistically significant contribution to the clusters. In addition to 12 significant variables in the K=5 analysis, the multi-birth variable becomes significant. Cluster 1 subjects (n=220) have high prevalence of OCD (p=2.0E^-38^), low ADHD (p=2.0E^-20^), higher prevalence of males (p=7.7E^-9^) and no trichotillomania (p=5.8E^-6^). Cluster 2 subjects (n=40) have trichotillomania (p=6.7E^-70^), with higher prevalence of subjects from the USA sites (p=6.9E^-4^) and OCD (p=8.0E^-4^). Cluster 3 subjects (n=176) have no OCD (p=9.9E^-90^), low prevalence of ADHD (p=4.2E^-20^), no trichotillomania (p=8.8E^-5^), higher prevalence of males (p=3.2E^-7^), and higher prevalence of Korean sites (p=6.2E^-05^). Cluster 4 subjects (n=98) have higher prevalence of females (p=1.2E^-39^), low ADHD (p=1.7E^-17^), high prevalence of OCD (p=1.1E^-15^), higher prevalence of multi-birth (p=1.5E^-7^), often later onset of tics (p=5.8E^-6^), and no trichotillomania (p=7.0E^-3^). Cluster 5 subjects (n=73) have no OCD (p=4.0E^-33^), higher prevalence of females (p=1.5E^-24^), no ADHD (p=1.9E^-16^), and often late onset of tics (p=7.5E^-10^). Cluster 6 subjects (n=181) have higher prevalence of ADHD (p=3.7E^-63^), higher prevalence of OCD (p=4.0E^-39^), higher prevalence of males (p=2.9E^-8^), and no trichotillomania (p=6.5E^-5^). Cluster 7 subjects (n=77) have no OCD (p=4.7E^-35^), high prevalence of ADHD (p=5.8E^-26^), all males (p=2.0E^-10^), and higher prevalence of ASD (p=8.8E^-3^).

Compared to K=5, K=7 splits subjects with OCD but not ADHD (K=5 cluster 3) and subjects without ADHD and OCD (K=5 cluster 4) into two clusters each based on sex, resulting in four smaller clusters (K=7 cluster 1, 4 and K=7 cluster 3, 5, respectively).

### BHC

BHC clustering using the data set with 14 categorical variables identified six clusters. We identified the variables that most significantly correlated (positively or negatively) with cluster identity (Figure 3A). Plotting cluster memberships along the top three PCs suggests robust clustering (Figures 3B-D) and similarity between clustering produced by K-Means and BHC. The detailed contribution and significance of each variable to each cluster are listed in Supplementary Data BHC.xlsx. Like the K-mean clustering analysis, 11 out of 14 variables show a statistically significant contribution to the clusters (Supplementary Data: BHC.xlsx – test.ch2 tab).

**Figure 3.**
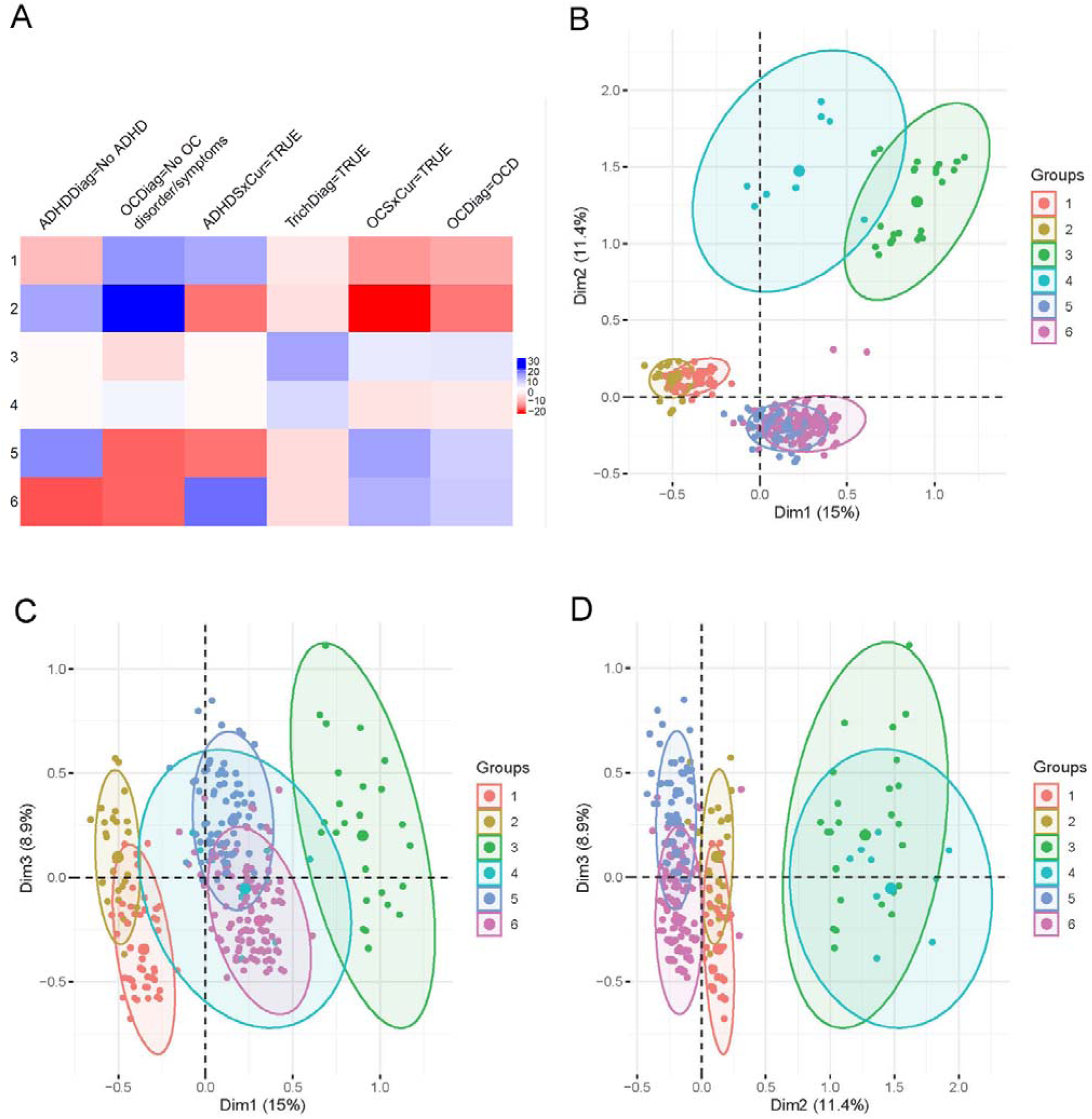
BHC Clusters. **A.** Major factors determining cluster characteristics based on V-test scores. Positive values are positive correlations, while negative values are negative correlations. The larger the value, the stronger the correlation. Only statistically significant variables are plotted. See Supplementary Table S2 for detailed variable descriptions. **B-D.** Cluster plots on first three PCs. Each subject is shown as a dot, while the color of the dot corresponding to the cluster assignment. The centroid of each cluster is shown as a big dot. Ovals indicate 95% normal confidence regions around clusters’ 2D point cloud.

Next, we examined the characteristics of each of the six clusters (Supplementary Table S9). Cluster 1 (n=120) comprises subjects with TD that have a high prevalence of ADHD (p=8E^-44^), no OCD (p=5.2E^-57^), and no trichotillomania (p=2.2E^-3^). Cluster 2 subjects (n=206) have low NDD comorbidities such as no OCD (p=7.9E^-110^), and ADHD (p=7.3E^-60^), an absence of trichotillomania (p=1.4E^-5^), and a low prevalence of ASD (p=3.6E^-5^). Cluster 3 subjects (n=30) have trichotillomania (p=2.9E^-47^), with a higher prevalence of OCD (p=2.8E^-8^), and are more likely to be from the USA (p=1.2E^-3^). Cluster 4 subjects (n=10) have trichotillomania (p=1.4E^-14^) and no OCD (p=1.3E^-3^). Cluster 5 subjects (n=250) have a high prevalence of OCD (p=2.1E^-49^), low levels of ADHD (p=2.3E^-64^), an absence of trichotillomania (p=8.1E^-7^), and an increased proportion of females (p=4.2E^-5^). Cluster 6 subjects (n=249) have ADHD (p=3.4E^-81^), OCD (p=1.3E^-42^), no trichotillomania (p=8.7E^-7^), and a higher proportion of males (p=3.6E^-5^).

### Comparison of K-Means and BHC’s results

K-Means uses a distance metric (Euclidean distance) and requires specification of the number of clusters. On the other hand, BHC uses an infinite mixture model to infer the optimal number of clusters and does not require specification of a distance metric. Having similar results from these two independent methods enhances our confidence in the results. Therefore, next we compared the cluster characteristics of K-Means and BHC clusters. Overall, K-Means K=5 clusters showed higher similarity to BHC clusters than K=7. Therefore, we focus on describing the comparison between K-Means K=5 and BHC clusters below.

Fraction of members in each K-Means cluster that are in each BHC cluster is depicted in a heatmap in Figure 4. Both methods resulted in four large clusters of similar sizes and characteristics, as illustrated in PC plots (Figures 2 and 3) and Table 1. These large clusters differ from each other only in minor characteristics and slightly varying memberships (Table 1). Besides the four large clusters, K-Means clustered all trichotillomania patients together in one cluster (cluster 2, Supplementary Table S7), while BHC split them into two clusters (with OCD (cluster 3, n=30) and without OCD (cluster 4, n=10), Supplementary Table S9). Given only 4.6% of the subjects (40 of 865) have trichotillomania and only 10 of those do not have OCD, we elect to follow the K-Means clustering of all trichotillomania patients and define five phenotypic clusters (subtypes) among the cohort. Most significant variables (low p-value) that characterize each subtype and differentiate it from others are highlighted in Table 1.

**Figure 4.**
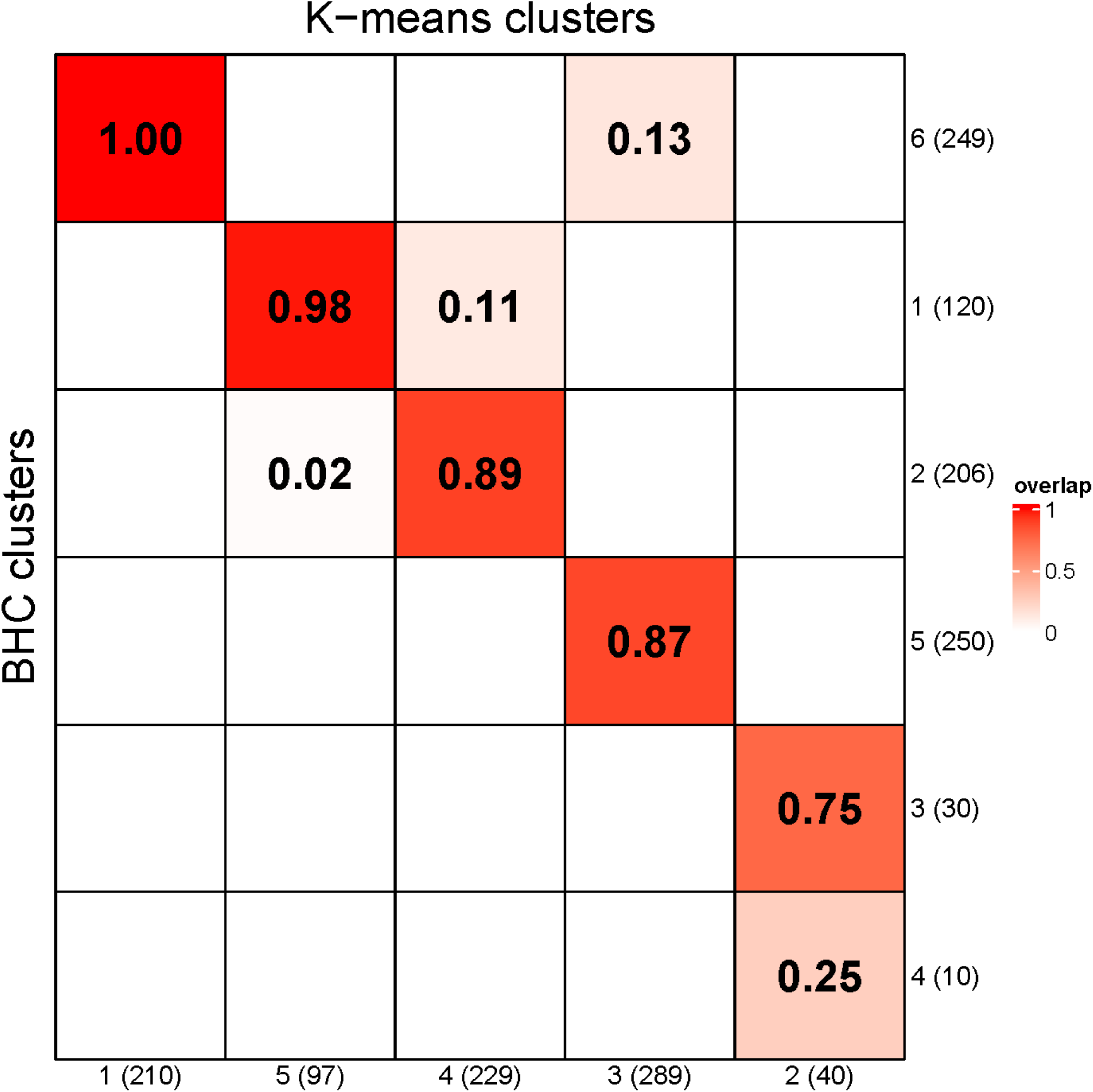
K-Means and BHC Cluster Comparison. Fraction of K-Means cluster members in each BHC Cluster in descending order of overlap. For example, 75% and 25% of the K-Means cluster 2 members are in BHC cluster 3 and 4, respectively.

**Table 1.** Comparison of K-Means (KM) and BHC Clusters.

| KM cluster | KM size+ | BHC cluster | BHC size* | Pos. Correlation | Neg. Correlation |
| --- | --- | --- | --- | --- | --- |
| 1 | 210 | 6 | 249 | **ADHD** | Trichotillomania |
|  |  |  |  | **OCD** |  |
|  |  |  |  | Male |  |
|  |  |  |  | ASD+ |  |
|  |  |  |  | USA+ |  |
|  |  |  |  | Tic Early Onset+ |  |
| 2 | 40 |  |  | **Trichotillomania** |  |
|  |  |  |  | USA |  |
|  |  | 3 | 30 | OCD+* |  |
|  |  | 4 | 10 |  | OCD* |
| 3 | 289 | 5 | 250 | **OCD** | **ADHD** |
|  |  |  |  | Female | Trichotillomania |
|  |  |  |  | Europe+ |  |
| 4 | 229 | 2 | 206 | South Korea+ | **OCD** |
|  |  |  |  | Tic Late Onset+ | **ADHD** |
|  |  |  |  |  | Trichotillomania |
|  |  |  |  |  | ASD |
| 5 | 97 | 1 | 120 | **ADHD** | **OCD** |
|  |  |  |  | Male+ | TicSxCur |
|  |  |  |  | ASD+ | Trichotillomania |

**Note:** Most statistically significant characteristics of each cluster are in bold
+ indicates of significance in K-Means
\* indicates of significance in BHC
V score with $p_{Adj} < 0.05$ is considered statistically significant (See Supplemental Data for details).

## Discussion

Tourette disorder has high phenotypic heterogeneity, high comorbidity with other NDDs, and varying rates of shared heritability with other NDDs, suggesting distinct underlying genetic causes for different TD subjects. In this study, we performed unsupervised clustering analysis of clinical data from 865 TD subjects in the TIC Genetics study. The analyses identified five phenotypic clusters and highlighted shared characteristics among TD patients. Prior attempts to identify specific genes or alleles contributing to disease risk have yielded only two genes with rare damaging variants that confer large effects and a single common variant of small individual effect [11, 39]. Stratified genetic analysis of TD subjects based on additional clinical features may help identify more genetically homogenous cohorts and improve power for gene discovery. Furthermore, identifying five phenotypic clusters could enable better diagnostics and treatment. Earlier attempts to identify subtypes often relied on small sample sets from single cohort [2, 23, 40, 41]. In addition, these studies usually relied on prior hypotheses regarding a subset of factors that were the cause of differentiation. To overcome these limitations, we performed unsupervised clustering analyses, using consistent clinical data from over 20 global sites, without filtering variables using prior hypothesis. This analysis enabled us to build on prior knowledge and identify novel TD subtypes.

A major challenge in clinical data analysis lies in the presence of noise and occasional misclassification, which can compromise the reliability of findings. Therefore, data selection and cleanup are essential. In addition, we also evaluated several unsupervised clustering methods to identify the optimal methods for our data set (Supplementary Table S3). After evaluating a variety of data set selection criteria and clustering methods, we selected only TD subjects for our final dataset (Supplementary Table S1) and identified two very different methods, K-Means and BHC, which worked well with our dataset. Final dataset was selected to be as complete and homogeneous as possible and the methods were selected to ensure stable clustering.

The two methods are quite different from each other. K-Means clustering was performed with PCs from MCA using the Euclidean Distance metric. Different metrics for assessing clustering quality generate different number of optimal clusters for our dataset. The number of clusters was ultimately chosen based on these metrics (Scree plot (Supplementary Figure S2) and cluster stability analysis (Supplementary Figure S3), Silhouette statistics (Supplementary Figure S4)) coupled with assessment of likely clinical relevance by experienced clinicians. On the other hand, BHC differs from traditional distance-based agglomerative clustering algorithms in several ways: (1) it defines a probabilistic model to compute the probability of a subject belonging to any of the existing clusters in the tree; (2) it uses a model-based criterion to decide on merging clusters rather than an ad-hoc distance metric; and (3) the number of clusters is automatically determined [34].

Because the primary goal of this study was to identify TD phenotypic clusters that are of high confidence as well as potentially clinically useful, we compared the clusters between the two methods and discussed the cluster characteristics with experienced clinicians to determine the clusters with the highest clinical relevance. In our analysis, K-Means identified five or seven clusters while BHC identified six clusters. In particular, the largest four clusters from K-Means K=5 and BHC methods have similar size and characteristics. The only difference between K-Means with k=5 and BHC was BHC split subjects with Trichotillomania (n=40) into those with OCD (n=30) and those without OC (n=10). As we started without any assumption regarding the number of clusters, the identification of similar sets of phenotypic clusters by two different methods (Table 1) increases our confidence in the results. After discussing with clinicians on our team with decades of experience, we decided five phenotypic clusters are the most clinically relevant results based on our analysis. In addition to confirming earlier findings on the importance of highly prevalent comorbidities such as ADHD and OCD [23], this large and diverse dataset identified new clusters that illustrate the importance of other clinically relevant factors such as trichotillomania and sex in TD subjects.

Trichotillomania is an often debilitating psychiatric condition characterized by recurrent hair-pulling, which can lead to disfiguring hair loss and significant distress and social impairment, and has been associated with elevated rates of anxiety, depression, ADHD, and OCD (DSM-5) [1]. In DSM-5, it is classified under OCD-related Disorders. The condition has significant heritability according to twin studies, a shared genetic liability with OCD and other OCD-related factors, as well as a distinct factor specific to hair-pulling and skin-picking [40]. Our study found subjects with TD and comorbid trichotillomania appear more likely to have OCD, reaffirming the earlier results.

Another advantage of the TIC Genetics cohort is its geographical diversity. Subjects were from the USA, Europe, Israel, and South Korea, which permitted us to assess the potential population-specific contribution to TD subtypes. In our analysis, regions are statistically significant factors in some clusters (e.g., K-Means K=5 cluster 1-4). However, their contributions to cluster characteristics are minimal (Supplementary Data: KMeans_3Dim_5Clusters.xlsx). We made some interesting observations among subjects from TIC Genetics centers in South Korea (69 Asians, ∼8.4% of the total cohort). Only about 10% of all Asian subjects with a diagnosis of TD also had a diagnosis of OCD, as compared to 40-60% observed in Caucasian subjects with TD diagnosis. A prior cross-national study also indicated a lower prevalence of OCD among Asians compared to the worldwide prevalence [42]. In addition, there are higher proportion of TD subjects with minimal or no NDD comorbidity from the USA and Europe than from South Korea. Although ancestry/genetic background could contribute to the observations, these apparent differences in rates of OCD and NDD comorbidity may also reflect the differences in ascertainment, referral patterns, cultural factors, diagnostic practices, and/or site-specific effects. Due to the uneven representation of different geographical regions in our study (e.g., ∼90% are Caucasians), confirming the population differences and determining the factors contributing to the differences are important next steps. Future studies with larger, more balanced sample collection and more data types (e.g., genetic data) are needed to confirm our observations and elucidate the mechanisms of the population differences.

While our dataset is larger than most prior studies and has more variables, this study still has several limitations. First, compared with phase 1, TIC Genetics phase 2 focused on recruiting simplex trios. However, we confirmed that this did not affect the unsupervised clustering because adding study phase as a variable did not affect the clustering results (data not shown). Second, as subjects with TD made up the vast majority of the subjects with tic disorders in the study, we only included subjects with TD to improve cluster stability. A larger dataset, including more subjects with other tic disorder diagnoses, could permit better clustering despite the noisiness of the data and provide better grouping of tic disorder subtypes in addition to TD. Third, our dataset might not be large or varied enough to identify all the underlying subtypes with high accuracy. Larger data sets will enable us to effectively use machine learning and other methods (e.g., Variational Auto Encoders) for unsupervised clustering. Lastly, in addition to observations by the clinicians, a portion of the data is self-reported by the subject or their parents and might produce an uncertain level of errors. Even though the clinicians in the TIC Genetics study were trained to record the symptoms and diagnosis in a consistent manner, there may still be undetected variations between individual clinicians and sites. Despite all these limitations, the large and diverse TIC Genetics cohort enabled us to enhance the current understanding by identifying new and potentially relevant factors and subtypes.

In the future, we will explore effective ways to include genetic data (e.g., SNP microarray and whole-exome sequencing data) available from the cohort to further identify TD subtypes. Another approach is to identify subtypes using genetic data and validate them using clinical data. The tools and methods we have evaluated and the workflow we developed here can also be used to evaluate hypotheses generated by domain experts. Ultimately, we plan to perform stratified analysis of genetic data based on the subtypes to identify new candidate genes and generate new insights into TD etiology, diagnosis, and treatment.

### Informed Consent and Institutional Review Board Statement

All adult participants and parents of children provided written informed consent along with written or oral assent of their participating child. The Institutional Review Board (IRB) of each participating site approved the study. All methods were performed in accordance with the relevant guidelines and regulations. All adult participants and parents of children provided written informed consent along with written or oral assent of their participating child. As a part of the TIC Genetics Study, this study was approved by the Institutional Review Board at Rutgers, State University of New Jersey.

### Consortia

Juliane Ball^51^, Leonie F. Becker^18,35^, Noa Benaroya-Milshtein^13,30^, Kate Bornais^15^, Danielle Cath^43,48^, Keun-Ah Cheon^59^, Barbara J. Coffey^60^, Lenrine Dalmeijer^39^, Erik M. Elster^12^, Dana Feldman^13,30^, Thomas V. Fernandez^57^, Carolin Fremer^8^, Lorena Garrote Espina^49,4^, Donald L. Gilbert^7^, Danea Glover^14^, Cristina Gómez Rapela^49^, Tammy Hedderly^27^, Gary A. Heiman^14^, Isobel Heyman^31^, Hyun Ju Hong^33^, Chaim Huijser^2,39^, Christina Kappler-Friedrichs^12^, Young Key Kim^23^, Young Shin Kim^53^, Robert A. King^58^, Nadine Kirchen^51^, Carolin Sophie Klages^8^, Yun-Joo Koh^38^, Florian Kraemer^51^, Subramanian Krishnamurthy^8^, Samuel Kuperman^20^, Bennett L. Leventhal^36^, Holan Liang^22,41^, Maria Loreta Lopez^60^, Marcos Madruga-Garrido^45,46^, Veronika Mailänder Zelger^51^, Osman Malik^28,47^, Marieke Messchendorp^52^, Malindi Mheen, van der^3,39,1^, Dararat Mingbunjerdsuk^54^, Pablo Mir^49,4^, Astrid Morer^11,34^, Kirsten Müller-Vahl^8^, Alexander Münchau^35^, Laura Muñoz-Delgado^49,4^, Tara L Murphy^31^, Cara Nasello^44,10,6^, Elena Ojeda-Lepe^49,4^, Veit Roessner^12^, Alyssa Rosen^24^, Guy Rouleau^16^, Simon Schmitt^8,21^, Chitra Shukla^24^, Sara Sopena^28^, Matthew W State^19^, Tamar Steinberg^13,30^, Frederika Tagwerker^51^, Zsanett Tarnok^56,26^, Joshua K. Thackray^44^, Meitar Timmor^13,30^, Jay A. Tischfield^14^, Max A. Tischfield^10,6^, Anne Uhlmann^12^, Ana Vigil-Pérez^11,34^, Frank Visscher^17^, Susanne Walitza^51^, Belinda Wang^19^, Jinchuan Xing^44^, and Samuel H. Zinner^55^.

^1^Amsterdam Public Health Research Institute, Amsterdam UMC, Amsterdam, the Netherlands

^2^Amsterdam UMC, Department of Child and Adolescent Psychiatry, Amsterdam, The Netherlands

^3^Amsterdam UMC, location University of Amsterdam, department of Child and Adolescent Psychiatry, Amsterdam, the Netherlands

^4^Centro de Investigación Biomédica en Red sobre Enfermedades Neurodegenerativas (CIBERNED), Instituto de Salud Carlos III, Madrid, Spain

^5^Centro de Investigacion en Red de Salud Mental (CIBERSAM), Instituto Carlos III, Spain

^6^Child Health Institute of New Jersey, Robert Wood Johnson Medical School, New Brunswick, NJ 08901

^7^Cincinnati Children’s Hospital Medical Center, Cincinnati, OH, USA

^8^Clinic of Psychiatry, Social Psychiatry and Psychotherapy, Hannover Medical School, Hannover, Germany

^9^Departamento de Medicina, Facultad de Medicina, Universidad de Sevilla, Seville, Spain

^10^Department of Cell Biology and Neuroscience, Rutgers University, Piscataway, NJ 08854

^11^Department of Child and Adolescent Psychiatry and Psychology, Institute of Neurosciences, Hospital Clinic Universitari, Barcelona, Spain

^12^Department of Child and Adolescent Psychiatry, Faculty of Medicine of the TU Dresden, Dresden, Germany

^13^Department of Child Psychiatry, Feinberg Child Study Center, Schneider Children’s Medical Center of Israel, Petach Tikva 4920235, Israel

^14^Department of Genetics and the Human Genetics Institute of New Jersey, Rutgers, the State University of New Jersey, Piscataway, NJ, 08854, USA

^15^Department of Human Genetics, McGill University

^16^Department of Neurology and Neurosurgery, Montreal Neurological Institute-Hospital, McGill University

^17^Department of Neurology, Van Weel Bethesda Ziekenhuis, Dirksland, The Netherlands

^18^Department of Pediatrics, University Hospital Medical Center Schleswig-Holstein, Campus Lübeck, Lübeck, Germany

^19^Department of Psychiatry and Behavioral Sciences, UCSF Weill Institute for Neurosciences, University of California, San Francisco, San Francisco, CA, USA.

^20^Department of Psychiatry, Professor of Child and Adolescent Psychiatry and Pediatrics, Emeritus MD, Carver College of Medicine, University of Iowa, Iowa City, IA, IA 52242, USA

^21^Department of Psychiatry, Psychotherapy and Psychosomatics, RWTH Aachen University, Aachen, Germany

^22^Department of Psychiatry, The University of Cambridge

^23^Department of Psychiatry, Yonsei Bom Clinic, Seoul, 03330, KR

^24^Division of Neonatology, Robert’s Center for Pediatric Research, Children’s Hospital of Philadelphia, Philadelphia, PA, USA.

^25^Division of Neurology, Children’s Hospital of Philadelphia, Philadelphia, PA, USA.

^26^Egyenlito Diagnostic, Behavioral Therapy and Consultation Centre in Budapest,

^27^Hungary Evelina London Children’s Hospital GSTT, Kings college London School of life sciences and medicine, London, UK

^28^Evelina London Children’s Hospital GSTT, Kings Health Partners AHSC, London, UK

^29^Evelina London Childrens Hospital, GSTT NHS Trust, Kings Health Partners AHSC, London,

^30^Gray Faculty of Medical and Health Sciences, Tel Aviv University, Tel Aviv 6997801, Israel

^31^Great Ormond Street Hospital for Children, and UCL Institute of Child Health, London, UK

^32^Great Ormond Street UCL Institute of Child Health, London, UK

^33^Hallym University Sacred Heart Hospital

^34^Institut d’Investigacions Biomediques August Pi i Sunyer (IDIBAPS), Barcelona, Spain Institute of Systems Motor Science, Center of Brain, Behavior and Metabolism.

^35^University of Lübeck, Lübeck, Germany

^36^Irving B. Harris Professor of Child and Adolescent Psychiatry, emeritus, The University of Chicago

^37^Korea Institute for Children’s Social Development and Rudolph Child Research Center, Seoul, South Korea

^38^Korea Institute for Children’s Social Development, Seoul, South Korea

^39^Levvel, Academic Center for Child and Adolescent Psychiatry, Amsterdam, the Netherlands

^40^National Health Insurance Service Ilsan Hospital, Goyang-si, South Korea

^41^North East London NHS Foundation Trust, UK

^42^Quantitative Biosciences Institute, University of California, San Francisco, San Francisco, CA, USA.

^43^Research department CMD MHC Drenthe, Assen, the Netherlands

^44^Rutgers, the State University of New Jersey, Department of Genetics and the Human Genetics Institute of New Jersey, Piscataway, NJ, USA

^45^Sección de Neuropediatría, Hospital Viamed Santa Angela de la Cruz. Seville.Spain

^46^Sección de Neuropediatría, Neurolinkia, Seville.Spain.

^47^South London and Maudsley NHS Foundation Trust.

^48^UMCG Groningen department of psychiatry & Rijksuniversity Groningen, the Netherlands

^49^Unidad de Trastornos del Movimiento, Servicio de Neurología, Instituto de Biomedicina de Sevilla (IBiS), Hospital Universitario Virgen del Rocío/CSIC/Universidad de Sevilla. Seville, Spain

^50^Universitat de Barcelona University Hospital of Psychiatry Zurich; Child and Adolescent Psychiaty and

^51^Psychotherapy, Neumuensterallee 9, 8032 Zurich, Switzerland

^52^University of Groningen, University Medical Center Groningen, Department of Child and Adolescent Psychiatry, Groningen, The Netherlands

^53^University of Texas Austin, Dell Medical School

^54^University of Washington and Seattle Children’s Hospital, Department of Neurology, Seattle, WA, USA

^55^University of Washington School of Medicine, Department of Pediatrics, Division of Developmental Medicine, 1925 NE Pacific Street, Box 356524, Seattle, WA 98195 USA

^56^Vadaskert Child and Adolescent Psychiatric Hospital, Budapest, Hungary

^57^Yale Child Study Center and Department of Psychiatry, Yale University School of Medicine, New Haven, CT, USA

^58^Yale Child Study Center, Yale University School of Medicine, New Haven, CT, 06510, USA

^59^Yonsei University College of Medicine, Severance Hospital, Seoul, South Korea

^60^Department of Psychiatry and Behavioral Sciences, University of Miami, Miller School of Medicine

## Supporting information

BHC Results

K-Means Results k=5

K-Means Results k=7

## Acknowledgments

We thank the families who have participated in and contributed to this study. Bio samples are available through the NIMH Repository and Genomics Resource (U24MH068457 to J.A.T.). We gratefully acknowledge the contributions of Mayra Aldecoa, Diana Bok, Maria Cruz, Kayla Delapenha, Kristen Fleming, Laura Ibanez-Gomez, Jessica D. Leuchter, Adam Lombroso, Daniela Martinez, and Vessela Zaharieva to the TIC Genetics study. We are also grateful to the NJCTS for facilitating the inception and organization of the TIC Genetics study.

## Authors’ Roles

Conceptualization, K.S., R.A.K., G.A.H., and J.X.; methodology, K.S and J.X.; formal analysis, K.S.; writing— original draft preparation, K.S.; editing, supervision, R.A.K., J.A.T., G.A.H., and J.X. All authors have read and agreed to the published version of the manuscript.

## Supplementary Tables

**File:** TD_SubTypes_SupplementaryTables.xlsx

**Supplementary Table S1.** TIC Genetics clinical data

**Supplementary Table S2.** Variables used for clustering

**Supplementary Table S3.** Unsupervised clustering methods tested

**Supplementary Table S4.** R packages used in this study

**Supplementary Table S5.** Different data sets evaluated for identifying subtypes

**Supplementary Table S6.** Component Weights by Sample (after MCA)

**Supplementary Table S7.** K-Means Cluster Size and Characteristics at K=5

**Supplementary Table S8.** K-Means Cluster Size and Characteristics at K=7

**Supplementary Table S9.** BHC Cluster Sizes and Characteristics

## Supplementary Data

**KMeans_3Dim_5Clusters.xlsx –** Category Description – Complete Results – k=5

**KMeans_3Dim_7Clusters.xlsx** – Category Description – Complete Results – k=7

**BHC.xlsx –** Category Description – Complete Results

High or low prevalence of a variable represents a statistically significant difference between actual distribution phenotype in a cluster and compared to the expected distribution based on random sampling of our selected data set. R FactoMiner package [43] was used to generate the cluster characteristics and variable importance.

## Supplementary Figures

**Supplementary Figure S1.**
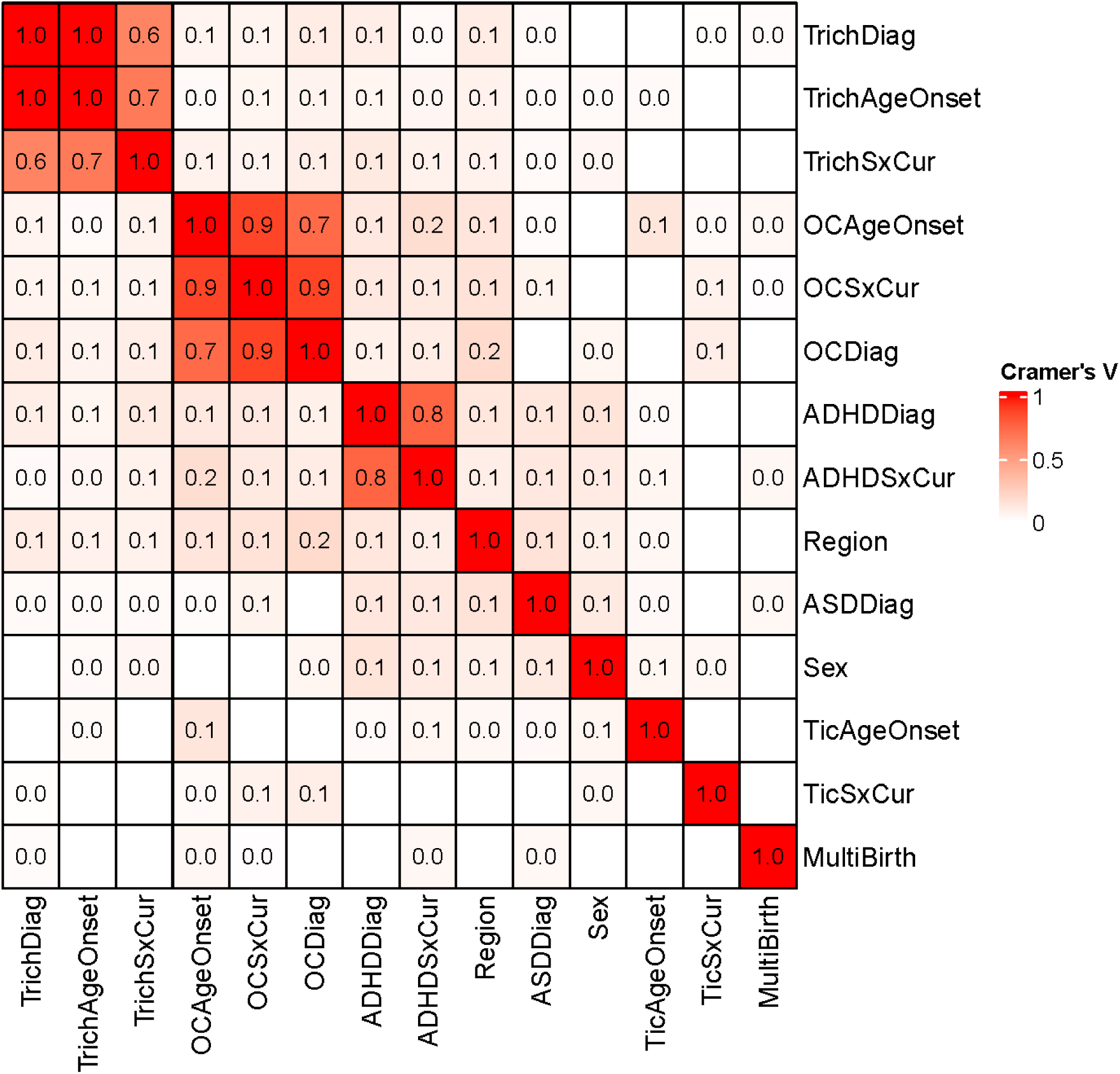
Pairwise variable association. Cramer’s V associations are shown, and empty cells (white) indicate no association.

**Supplementary Figure S2.**
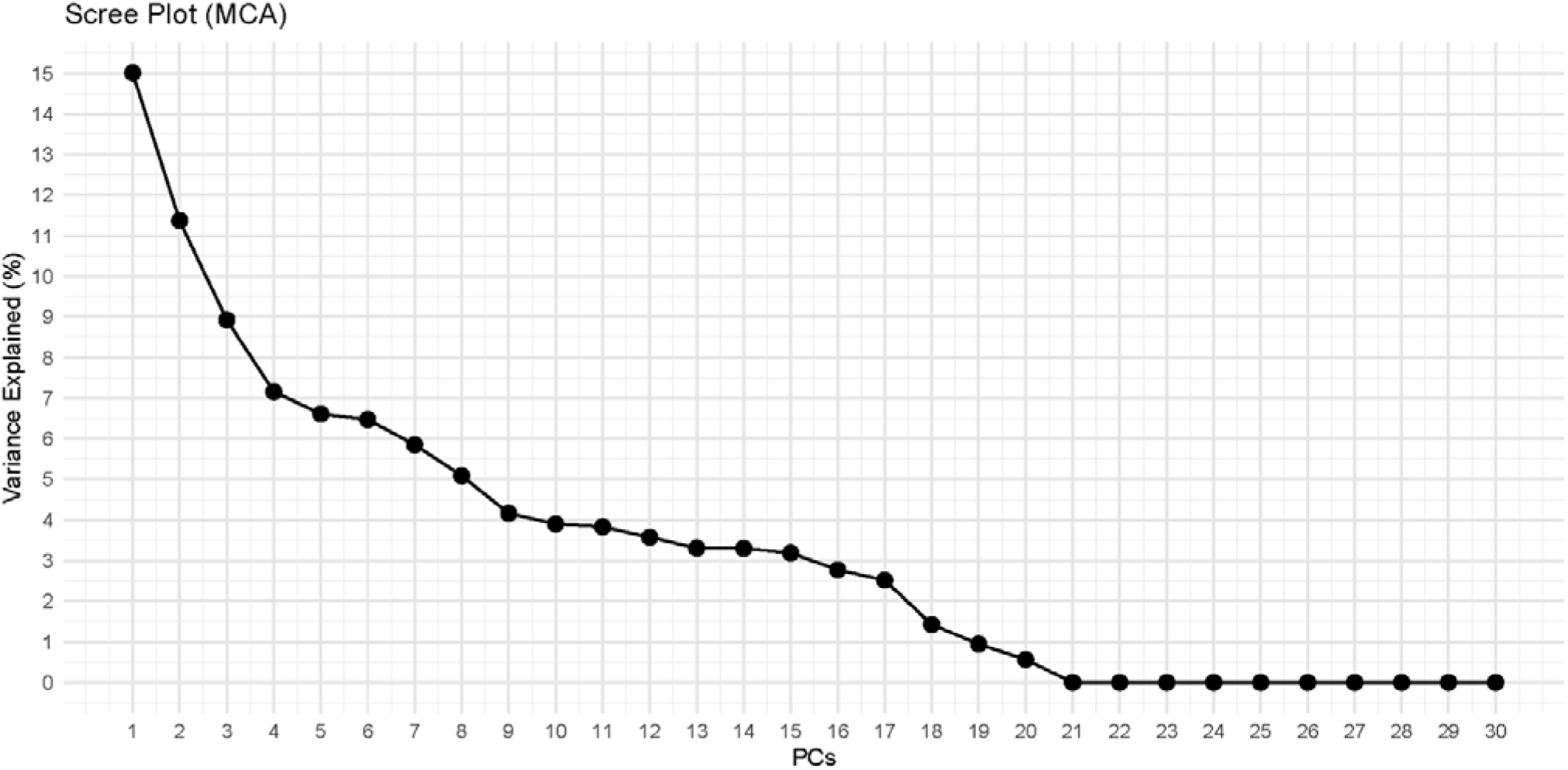
Scree plot of the percentage of variance explained by each PC in MCA.

**Supplementary Figure S3.**
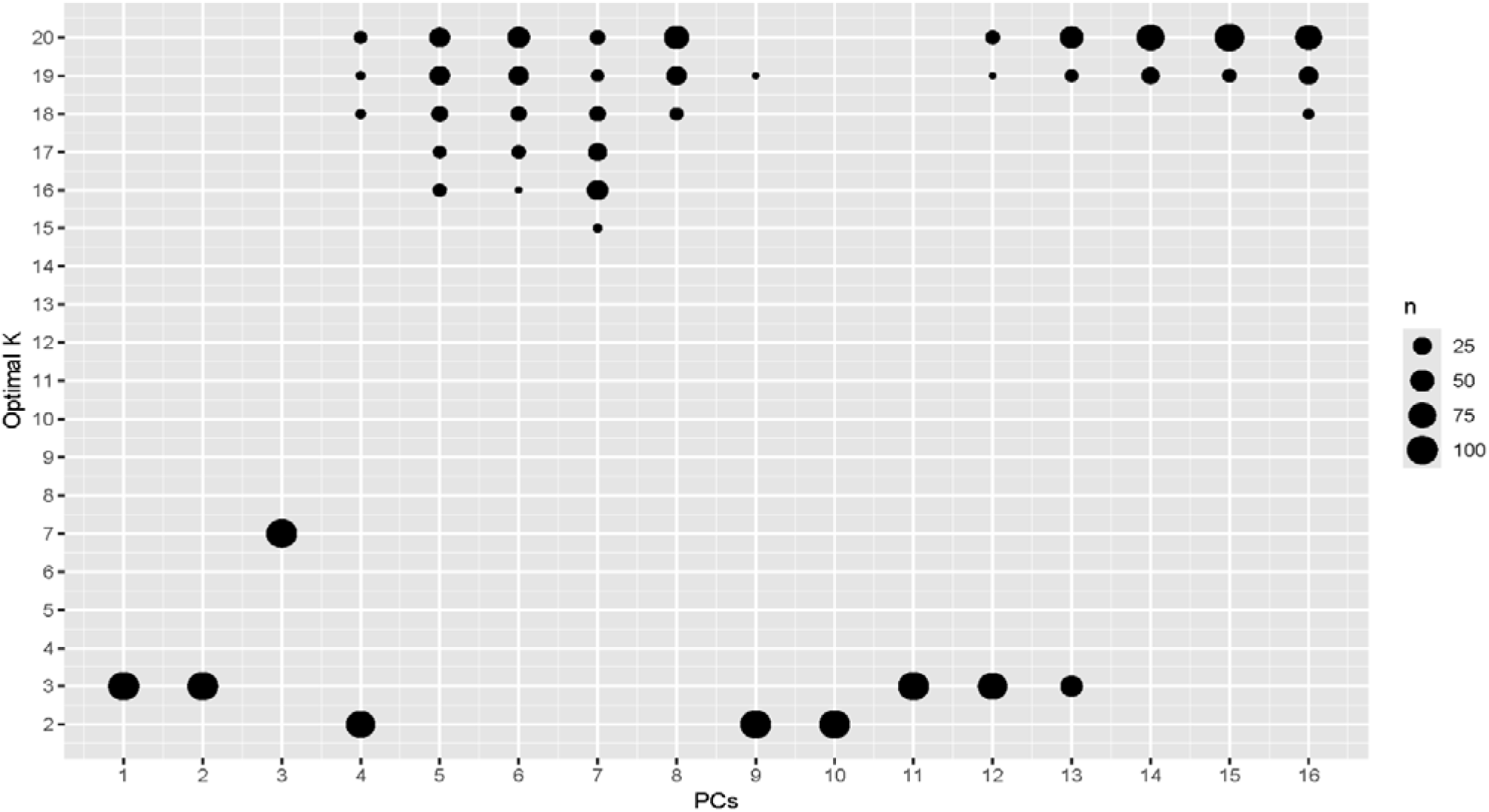
Cluster stability analysis for K-Means with k=2 to 20 and PCs 1-16. K-Means cluster analysis was repeated 100 times for the stability analysis and the size of the dot shows the number of repeats with an optimal K.

**Supplementary Figure S4.**
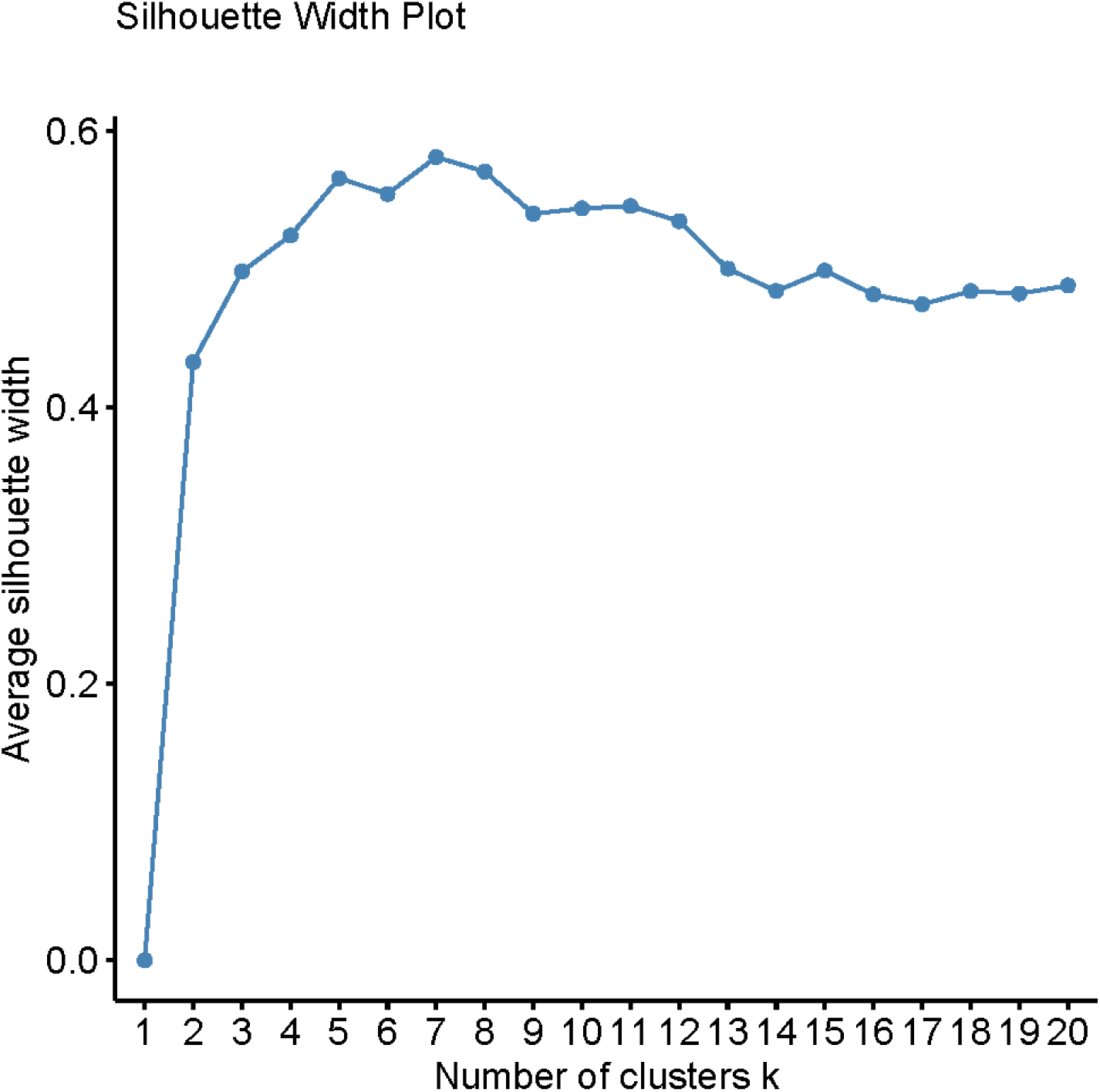
Silhouette Plot for K-Means analysis with k=1 to 20.

